# Acute plasma VEGF-A levels are associated with raised intracranial pressure and chronic lesion volume after Traumatic Brain Injury

**DOI:** 10.1101/2025.02.17.25322293

**Authors:** Lucia M. Li, Eleftheria Kodosaki, Amanda Heslegrave, Eyal Soreq, Giovanni Nattino, Elena Garbero, Karl A. Zimmerman, Neil S.N. Graham, Federico Moro, Deborah Novelli, Primoz Gradisek, Sandra Magnoni, Henrik Zetterberg, Guido Bertolini, David J Sharp

## Abstract

**Background and Objectives:** Severe Traumatic Brain Injury (TBI) is associated with secondary injury and poor outcomes, but the underlying mechanisms are poorly understood. Vascular mechanisms may be important. We aimed to characterise how blood vascular endothelial growth factor A (VEGF-A) levels are affected by TBI, and its associations with secondary injury and functional outcome.

**Methods:** We retrospectively analysed data from two multi-centre, international, prospective observational studies (CREACTIVE and BIO-AX-TBI) with follow-up of up to 1 year. These cohorts comprised adults with moderate-severe TBI (Mayo classification), recruited on admission to hospital (BIO-AX-TBI) and the intensive care unit (ICU) (CREACTIVE). Controls included non-TBI trauma (NTT) and uninjured adults. Plasma VEGF-A levels and TBI biomarkers (Neurofilament light [NFL], glial fibrillary acidic protein [GFAP], total Tau, UCH-L1, S100B) were measured on ICU admission and ∼5 days later (CREACTIVE), or at 5 timepoints from admission to 12 months post-TBI (BIO-AX-TBI), and compared to NTT and control groups. In BIO-AX-TBI, MRI assessment was performed at subacute and chronic timepoints. Functional outcomes (Glasgow Outcome Scale-Extended) were measured at 6 and 12 months. Plasma VEGF-A was measured using the OLINK® Target 96 Inflammatory platform, which reports in arbitrary standardised units (NPX), and TBI biomarkers were measured using Simoa® or Millipore platforms.

**Results:** Data was available from 195 TBI (21% female, mean age 45.30years), 24 NTT (8%, 43.98) and 89 CON (44%, 42.39) in BIO-AX-TBI, and 1146 TBI (25%, 56.29) in CREACTIVE. Plasma VEGF-A was elevated acutely after both TBI (estimated mean difference=0.45NPX, SE=0.09, p<0.001) and NTT (estimated mean difference=0.74NPX, SE=0.16, p<0.001), but remained raised after the initial timepoint only in TBI patients, peaking at day 16. Higher acute VEGF-A was associated with increased odds of refractory raised intracranial pressure (r-rICP) (maximum Odds Ratio for r-rICP=1.69, p=0.031), higher lesion volume (estimated increased lesion volume=20.14ml, SE=8.20ml, p=0.02), and worse functional outcomes (maximum Odds Ratio for worse outcome category=2.51, p<0.001).

**Discussion:** There is a sustained rise in plasma VEGF-A after TBI, which is associated with r-rICP and chronic injury markers, suggesting vascular pathophysiology is important after TBI. Further research is needed to explore mechanisms.

## Introduction

Traumatic Brain Injury (TBI) results in heterogenous clinical outcomes, reflecting both initial damage and secondary injury like cerebral oedema and raised intracranial pressure (rICP)^1^. Understanding the pathophysiology of secondary injury can guide therapeutics development.

Vascular endothelial growth factor A (VEGF-A), produced by almost all brain cell types, has multiple roles in the brain including neuronal survival, blood-brain barrier (BBB) integrity, and peripheral inflammatory cell recruitment^2^. In TBI models, glial cell VEGF-A contributes to BBB breakdown^3^, whilst VEGF-A inhibition reduces BBB permeability through modulation of tight junction proteins and cytokines^4,5^. BBB breakdown leads to cerebral oedema and rICP, which is associated with worse TBI outcomes^6^. Elevated VEGF-A levels post-TBI have been reported in plasma, microdialysate and peri-contusional tissue^7–10^, correlating with worse outcomes^7^. However, the link between plasma VEGF-A levels and rICP has not been previously investigated. Other important questions include the trajectory and specificity of post-TBI plasma VEGF-A elevation, and its association with injury factors.

We investigated acute plasma VEGF-A levels and its clinical associations in a multi-centre European TBI cohort, the CREACTIVE study (1146 patients), replicating our analyses in separate cohort, the BIO-AX-TBI study (195 patients)^11–13^. Both cohorts had clinical assessments, follow-up data, and acute plasma VEGF-A, neuronal and astroglial biomarker levels. These biomarkers included: NFL (neurofilament light), GFAP (glial fibrillary acidic protein), total Tau, UCH-L1 (ubiquitin C-terminal hydrolase 1) and in BIO-AX-TBI, S100B (S100 calcium binding protein B)^14–17^. Acute GFAP levels reflect extent of intracranial injury, showing strong associations with clinical severity, medical care requirement and traumatic lesions visible on CT and MRI^18,19^. Chronic plasma NFL, Tau, and GFAP may reflect ongoing neuronal injury^15,20,21^. The BIO-AX-TBI cohort additionally provided later blood samples, longitudinal MRI scans, and both non-injured (CON) and non-TBI trauma (NTT) control groups.

We hypothesised that plasma VEGF-A is elevated acutely after TBI, and higher than in non-TBI trauma. We also expected plasma VEGF-A to be associated with risk of rICP, independently of the extent of initial injury, and also with injury severity, other TBI biomarkers, and functional outcomes.

## Methods

### Participant cohort

Data was available from 1146 TBI participants admitted into the intensive care unit (ICU) in the CREACTIVE study. Participants had blood samples upon ICU admission (“*acute 1*”) and ∼5 days after (“*acute 2*”). Functional outcomes were assessed at 6 months post-TBI with the Glasgow Outcome Scale-Extended (GOS-E)^22^ (FIG 1).

**Figure 1:**
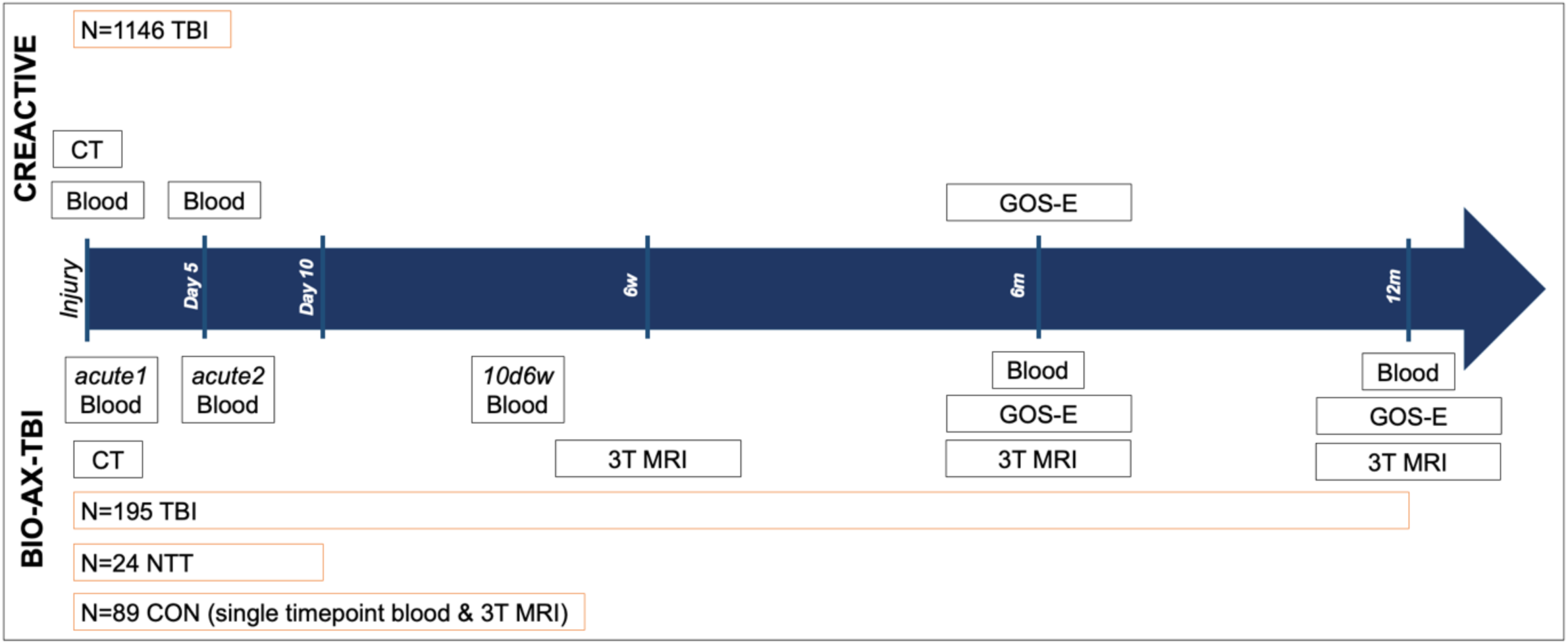
schematic summarising the data collection timings from each cohort. TBI=traumatic brain injury; NTT=non-TBI trauma; CON=non-injured controls; CT=computed tomography scan; CSF=cerebrospinal fluid; GOS-E=Glasgow Outcome Scale-Extended; MRI=magnetic resonance imaging; 10d6w=10 days to 6 weeks

Data was also available from 195 TBI, 24 non-TBI trauma (NTT, 46% limb fractures), both recruited at point of hospital admission, and 89 non-injured control (CON) participants from the BIO-AX-TBI study. Blood samples were taken at two time points up to 10 days post injury (“*acute 1*” and “*acute 2*”). TBI participants additionally had bloods and 3T MRI performed between 10 days and 6 weeks of injury (subacute, “*10d6w*”), at 6 and 12 months (chronic, “*6m*” and “*12m*”). GOS-E was collected at 6m and 12m (FIG 1).

Key demographic and clinical information for all cohorts are summarised in Table 1 (full details previously published^11–13,23,24^). Written informed consent or assent (for participants without capacity) was obtained for all participants, and local ethics boards approved the studies.

**Table 1:**
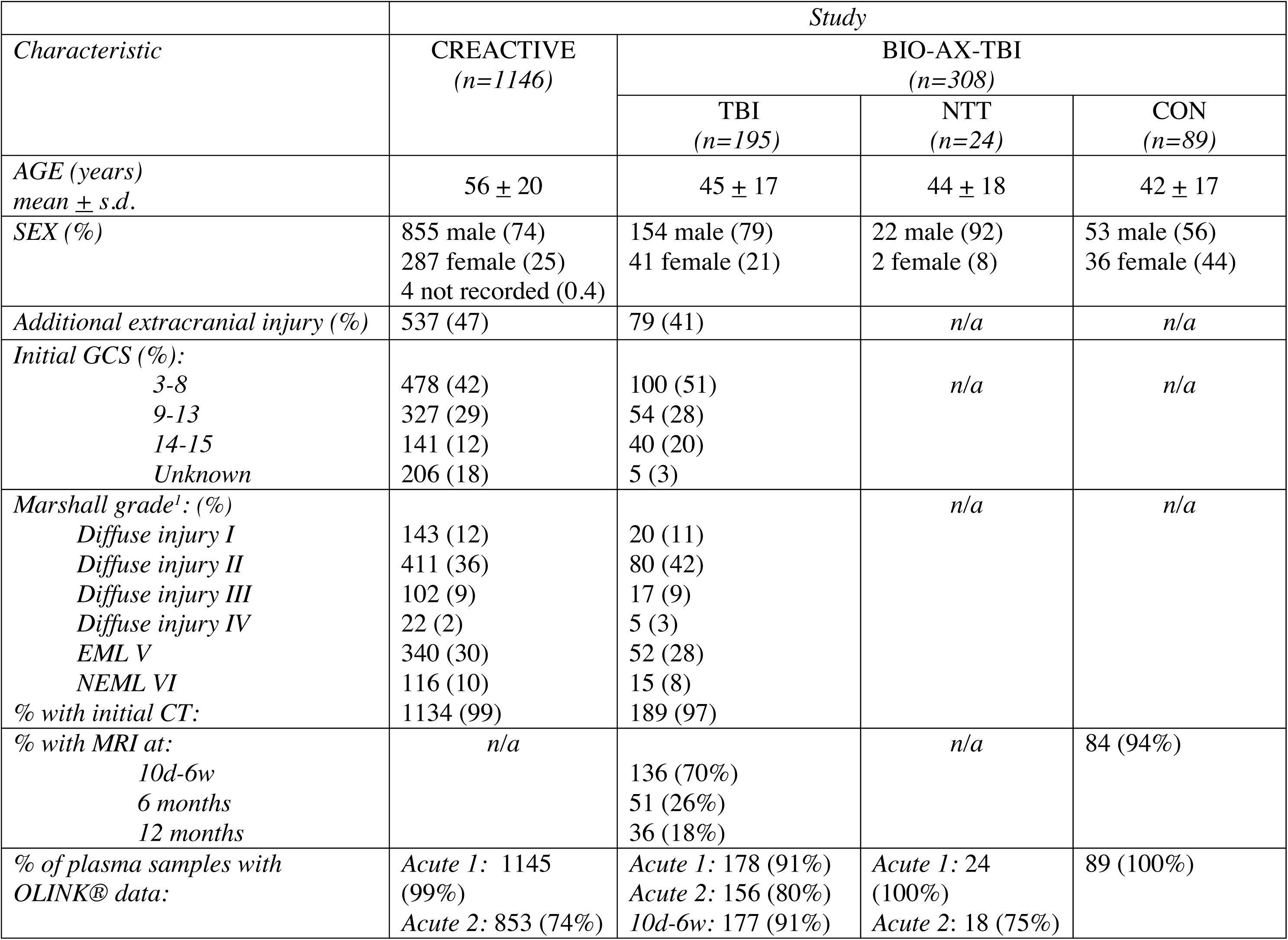
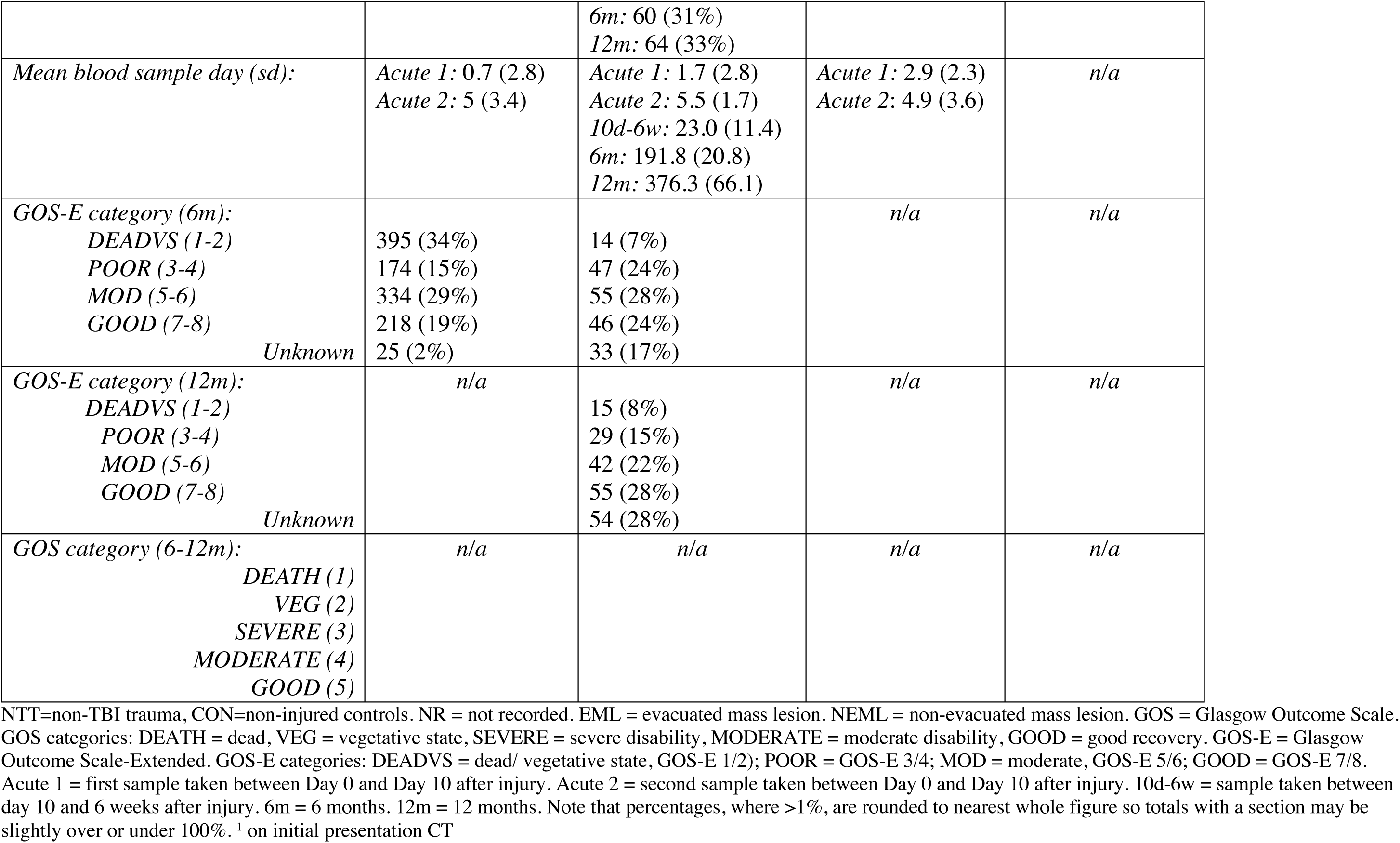
Demographic, Baseline Clinical Characteristics and Outcomes in the CREACTIVE and BIO-AX-TBI study.

### MRI Acquisition and Processing

Full protocol details for MRI acquisition, preprocessing and analysis are previously described^13,14^. Briefly, 1.5T (Florence and Niguarda sites) or 3T (all other sites) MRI was acquired at *10d6w*, *6m* and *12m* timepoints. Change in white and grey matter volumes were calculated as the Jacobian Determinant (JD) between the *10d6w* and *6m* scans, and between the 6m and 12m scans. Lesion volume was calculated from manually drawn lesion masks (by NG, a neurologist, and KZ, FM, experienced neuroimaging researchers), using T1w and T2 FLAIR scans, and volumes extracted from the masks with *fslstats* from the FSL imaging analysis software package^25^. White matter injury was assessed with diffusion tensor imaging. After registration of diffusion scans to DTITK space^26^ and generating voxelwise maps of fractional anisotropy (FA) using a tract-based spatial statistics approach, we calculated the z-scored mean FA across the whole skeletonised white matter by comparing patients recruited on a specific scanner to values from controls acquired on the same scanner. Mean z-scored FA was then extracted for the corpus callosum and whole white matter skeleton.

### Blood Sample Processing

Blood was collected in EDTA and centrifuged for plasma collection, which was frozen immediately and stored until use. Full details are previously described (CREACTIVE^11^ and BIO-AX-TBI^14^). Plasma levels of 92 inflammation markers were measured on the OLINK® Signature Q100 platform (OLINK® Proteomics AB, Uppsala, Sweden) using the “Target 96 Inflammation” panel. This panel uses Proximity Extension Assay (PEA) technology^27^. CREACTIVE samples had been previously tested by the Uppsala Analysis Service at OLINK® Proteomics. Subsequently, the UCL Biomarker Factory analysed all available BIO-AX-TBI samples. OLINK® biomarker data is normalised to minimise variation, and are reported as normalised protein expression (NPX) values, an arbitrary unit on a log2 scale. An increase of 1 NPX equates to a doubling of concentration. NFL, GFAP, total tau (TAU) and UCH-L1 was measured in CREACTIVE samples with the Quanterix Simoa® Neuro 4-PLEX-B assay. BIO-AX-TBI samples were analysed on a Simoa®-HD1 platform for GFAP, TAU, NFL and UCH-L1, and with a Millipore enzyme-linked immunosorbent assay kit for S100B^14^. We used *acute 1* plasma GFAP levels as a quantitative measure of initial intracranial injury extent.

### Statistical Analyses

Analyses were carried out with R (version 4.3.2) in RStudio (2021). For CREACTIVE data, we used linear mixed effects (LME) models to investigate the effect of Glasgow Coma Scale (GCS) and time on VEGF-A levels, with age and additional extracranial injury (ECI) as covariates. For the BIO-AX-TBI study, due to the availability of control groups, we used LME models to determine the impact of time and group on VEGF-A levels, with age as a covariate. Post-hoc t-tests identified the significant pairwise comparisons in the models, with FDR correction for multiple comparisons. We estimated the day of peak post-TBI plasma VEGF-A expression using all available *acute 1*, *acute 2* and *10d6w* TBI sample data from BIO-AX-TBI. We fitted a mixed effects model with quadratic polynomial transformation of the post-injury day data.

We used Spearman’s correlations (FDR corrected for multiple comparisons) to investigate the relationship between plasma levels of VEGF-A and neuronal/ astroglial TBI markers including NFL, GFAP, TAU, UCH-L1 and S100B. Further detail in *Supplementary Methods*.

We used logistic regression (*polr*) to investigate the association of VEGF-A levels with presence of intracranial blood (ICH, defined as intraparenchymal, subdural, subarachnoid or intraventricular) and presence of raised intracranial pressure (rICP) and refractory raised ICP (r-rICP, defined as persistent ICP>30mmHg despite standard ICP therapies). We included age and *acute 1* plasma GFAP level, an indicator of extent of initial intracranial injury^18^, as covariates. GFAP level was log2 transformed to enable comparable interpretation with VEGF-A NPX units. Further detail in *Supplementary Methods*.

We used linear models to investigate the association of peak VEGF-A levels with MRI measures of injury (lesion volume and white matter fractional anisotropy [FA]) and ongoing neurodegeneration (change in white matter and grey matter volumes between MRI scans), in the BIO-AX-TBI cohort. Further detail in *Supplementary Methods*.

Functional outcome was assessed with the Glasgow Outcome Scale Extended (GOS-E), categorised as “GOOD” for scores 7-8, “MOD” (moderate) for scores 5-6, “POOR” for scores 3-4, and “DEADVS” for scores 1-2. We used a Kruskal-Wallis test, with 2-tailed post-hoc Dunn’s test to identify significant post-hoc comparisons of peak plasma VEGF-A between GOS-E categories. We used logistic regressions (*polr*) to investigate the relative contribution of VEGF-A levels to GOS-E, with age and *acute 1* GFAP (log2 transformed) as covariates. Full model details, including the numbers and demographics of TBI patients going into each analysis, based on data availability, are summarised in *Supplementary Methods* and SI TABLE3.

We split the BIO-AX-TBI TBI participants into 4 groups depending on VEGF-A trajectory in the acute-subacute period. We tested for differences in characteristics and outcomes between different trajectory groups using Chi-squared tests for proportions, one-way ANOVAs for continuous variables and logistic regression (*polr*) for outcomes requiring adjustment for covariates.

#### Data Availability Statement

the datasheets, R workspace and R code scripts will be made available to any reasonable request, and subject to data sharing agreements.

## RESULTS

### Elevated plasma VEGF-A after TBI sustained into the subacute period

Plasma VEGF-A levels in the CREACTIVE study showed a significant rise between the *acute 1* and *acute 2* timepoints (t = −11.781, df = 1881.5, p-value<0.0001) (FIG 2A).

**Figure 2:**
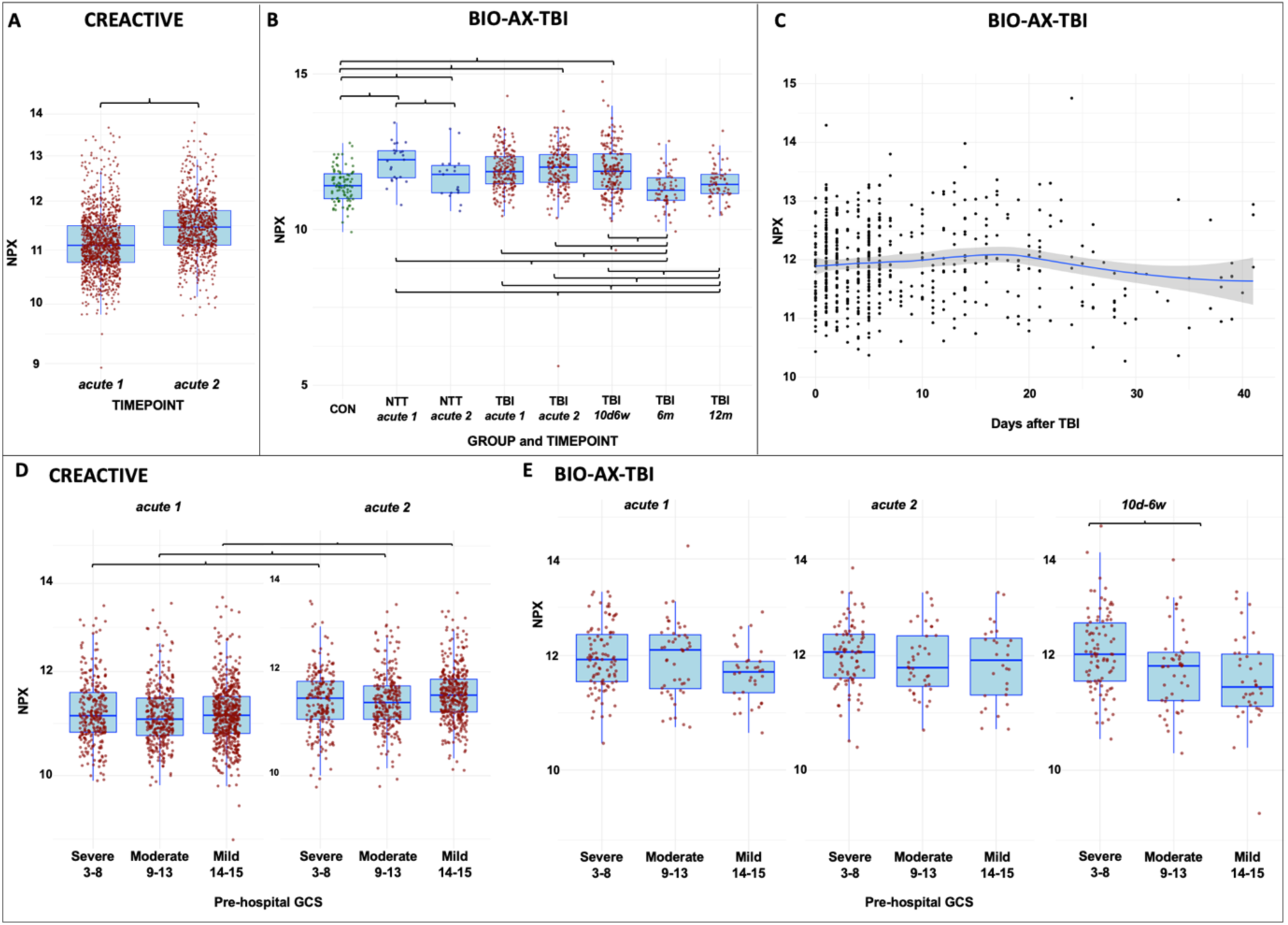
(A) Boxplot illustrating plasma VEGF-A between the D0 and D5 timepoints in the CREACTIVE cohort (bracket indicating significant difference). (B) Boxplot illustrating plasma VEGF-A levels in non-injured controls (CON), non-TBI trauma controls (NTT) and TBI participants at acute 1 (first acute blood sample in the first 10 days of injury), acute 2 (second acute blood sample in the first 10 days of injury), 10 days to 6 weeks after TBI (10d-6w) and at 6 months (6m) and 12 months (12m) after TBI (brackets indicate significant FDR-corrected differences). (C) Trajectory of plasma VEGF-A in the BIO-AX-TBI cohort, with the peak at day 16 after TBI, visualised with LOESS curve. (D) Boxplot illustrating the statistically significant rise in plasma VEGF-A between the D0 and D5 timepoints, irrespective of the pre-hospital GCS category, in the CREACTIVE cohort. (E) Boxplots illustrating plasma VEGF-A levels at different timepoints for patients with different pre-hospital GCS in the BIO-AX-TBI cohort. Brackets indicate statistically significant comparisons in post-hoc testing.

In the BIO-AX-TBI study, there was a significant effect of Group (F(2,525.69)=6.37, p=0.0018) and Timepoint (F(4,604.14)=12.33, p=1.20×10^-9^). Plasma VEGF-A levels were raised in both acute TBI and NTT (*acute 1* timepoint), compared to non-injured controls (CON). Thus, the acute rise in plasma VEGF-A was not TBI-specific. However, NTT plasma VEGF-A levels were comparable with CON *acute 2*, whereas TBI plasma VEGF-A levels remained elevated into the subacute (*10d6w*) period (FIG 2B). The peak plasma VEGF-A was estimated at 16 days post-TBI (FIG 2C).

### Acute plasma VEGF-A is associated with age, extracranial injury and clinical injury severity

In the CREACTIVE data, there were significant effects of age (F(1,1989)=114.47, p<0.0001) and presence of extracranial injury (ECI) (F(1,1989)=12.06, p=0.0005) on plasma VEGF-A levels. Older patients (*t*=9.74) and patients with ECI (*t*=2.91) had higher levels (SI FIG 1). There was no effect of clinical injury severity, as assessed by pre-hospital GCS (Glasgow Coma Scale) category (FIG 2D).

In contrast, the BIO-AX-TBI data showed a significant effect of pre-hospital GCS (F(2,499)=8.94, p=0.0002) on plasma VEGF-A levels (FIG 2E). Patients with a pre-hospital GCS of 3-8 had higher plasma VEGF-A levels than those with GCS 14-15 (*t*=4.01). There was no main effect of timepoint, nor any interaction between timepoint and pre-hospital GCS. As for CREACTIVE, there were significant effects of age (F(1,497)=18.34, p<0.0001) and presence of ECI (F(1,497)=11.92, p=0.0006), with older patients (*t*=4.22) and patients with ECI (*t*=2.94) having higher plasma VEGF-A levels (SI FIG 2).

### Early plasma VEGF-A levels correlate with early and late neuronal and astroglial blood biomarkers

In line with our previous work^14^, we show that GFAP, total Tau, UCH-L1 and S100B plasma levels are increased at *acute 1* and then fall, whereas plasma NFL levels rise into the subacute (*10d6w*) period (SI FIG 3A, C). There were multiple positive correlations between acute, subacute and peak plasma VEGF-A and neuronal/ astroglial biomarker levels (SI FIG 3B, D). The strongest relationships were seen between subacute VEGF-A and UCH-L1 (r_s_=0.43), peak VEGF-A and S100B (r_s_=0.41), and peak VEGF-A and 6m NFL (r_s_=0.41, all FDR-corrected p<0.05). Full results are detailed in Supplementary Results.

### Higher acute plasma VEGF-A increases risk of having refractory raised intracranial pressure

VEGF-A may contribute to raised intracranial pressure (rICP) and refractory raised intracranial pressure (r-rICP) through increasing BBB breakdown^4,5^. Additionally, given high expression in vessel endothelium, blood vessel disruption accompanying intracranial haemorrhage may also result in elevated plasma VEGF-A^30^.

In the CREACTIVE data, *acute 2* plasma VEGF-A showed a significant positive association with having r-rICP, whilst *acute 1* plasma VEGF-A showed a significant negative association with having rICP (FIG 3A,B). For a 1-unit increase in NPX (i.e. doubling) of *acute 1* VEGF-A, the odds of having rICP was 0.67 (p=0.021). For a 1-unit increase of *acute 2* VEGF-A, the odds of having r-rICP was 1.69 (p=0.031). In a partial replication in the BIO-AX-TBI data, *acute 2* plasma VEGF-A showed a significant positive association with having either r-rICP or having rICP (FIG 5D, E). For a 1-unit increase in NPX (i.e. doubling of plasma VEGF-A), the odds of having rICP was 2.25 (p=0.021), and of having r-rICP was 2.91 (p=0.026).

**Figure 3:**
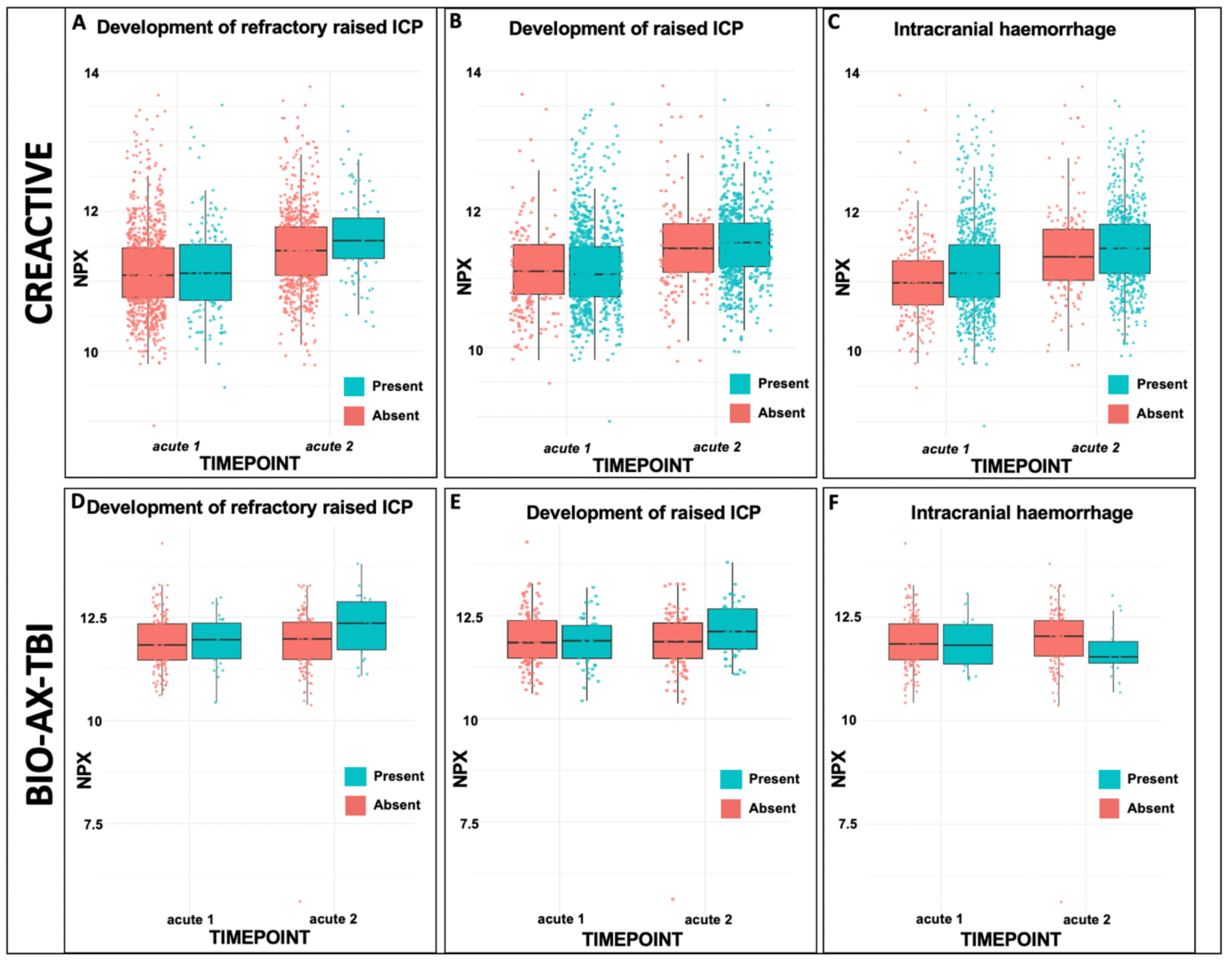
**(A-C) CREACTIVE cohort:** Boxplots illustrating relative plasma VEGF-A levels at acute 1 and acute 2 timepoints for (A) TBI patients who develop refractory raised ICP, (B) who develop raised ICP, and (C) who have intracranial haemorrhage on admission CT. **(E-F) BIO-AX-TBI cohort:** Boxplots illustrating relative plasma VEGF-A levels at acute 1 and acute 2 timepoints for (A) TBI patients who develop refractory raised ICP, (B) who develop raised ICP, and (C) who have intracranial haemorrhage on admission CT.

In both CREACTIVE and BIO-AX-TBI cohorts, there were also significant positive associations between *acute 1* plasma GFAP, indicative of intracranial injury extent, and occurrence of rICP or r-rICP. There were significant negative associations between age and occurrence of rICP and r-rICP. There were significant positive associations between age and *acute 1* GFAP, but not VEGF-A, with presence of ICH (FIG 3C, E). See Supplementary Results for full details.

### Peak plasma VEGF-A levels are associated with chronic lesion volumes

Peak plasma VEGF-A levels were positively associated with lesion volume at 12 months, accounting for age and *acute 1* GFAP level (coefficient=20,138.9, p=0.02) (SI FIG 4). For every unit increase (i.e. doubling) of peak plasma VEGF-A levels, the lesion volume at 12 months increased by 20,138.9mm^3^ (or 20.14ml). There were no other significant associations (see Supplementary Results).

### Acute plasma VEGF-A levels are higher in TBI patients with worse functional outcomes

In the CREACTIVE cohort, TBI patients with worse *6m* GOS-E had higher *acute 1* (X^2^(3)=73.25, p<0.0001) and *acute 2* (X^2^(3)=110.36, p<0.001) plasma VEGF-A levels (FIG 4A, 4B). These associations were significant even after accounting for age and *acute 1* plasma GFAP, as an indicator of intracranial injury. The odds ratio (OR) of having a worse GOS-E category with a doubling of *acute 1* VEGF-A levels was 1.72 (p<0.0001), and 2.51 (p<0.001) with a doubling of *acute 2* VEGF-A. Further detail in Supplementary Results.

**Figure 4:**
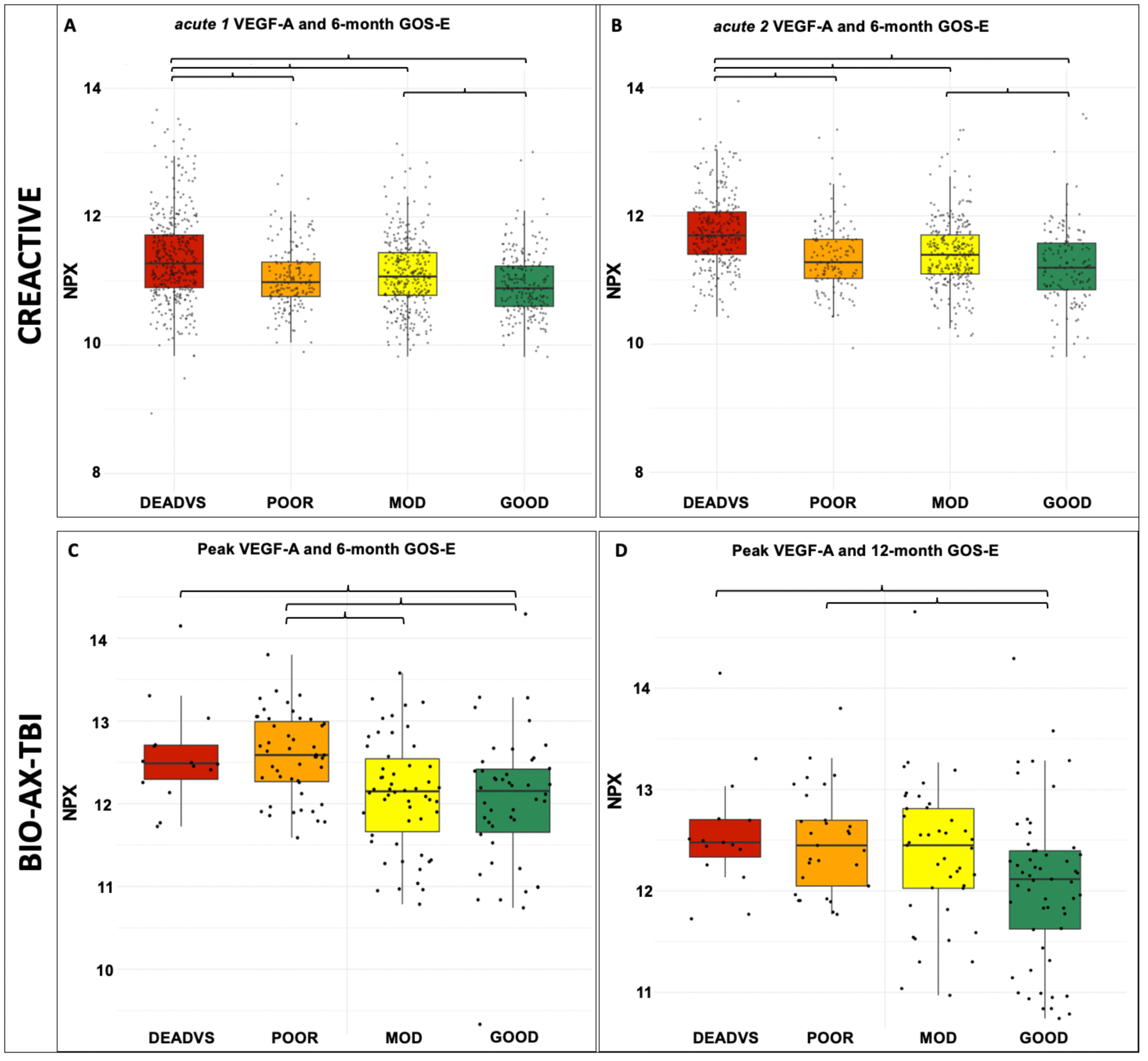
Boxplots showing plasma VEGF-A levels and the relationship to Glasgow Outcome Scale Extended (GOS-E) assessment of functional outcome. (A) D0 VEGF-A levels and 6 month GOS-E in CREACTIVE cohort. (B) D5 VEGF-A levels and 6 month GOS-E in CREACTIVE cohort. (C) Peak VEGF-A levels and 6 month GOS-E in BIO-AX-TBI cohort. (D) Peak VEGF-A levels and 12 month GOS-E in BIO-AX-TBI cohort. Brackets indicate statistically significant post-hoc pairwise comparisons, reported in SI TABLE4. DEADVS = dead/ vegetative state; MOD=moderate outcome.

Partially replicating these results in the BIO-AX-TBI cohort, peak plasma VEGF-A was higher in TBI patients with worse *6m* GOS-E at 6 months (X²(3)=19.82, p=0.0002) (FIG 4C) and 12 months (X^2^(3)=11.95, p=0.0076) (FIG 4D). A doubling of peak VEGF-A levels corresponded with an increased odds of moving into a worse GOS-E category at 6 months (OR=2.20, *p=0.0002*) and at 12 months (OR=2.06, *p=0.002*) (SI TABLE4). However, after additionally accounting for *acute 1* GFAP, peak VEGF-A plasma levels were no longer significantly associated with *6m* and *12m* GOS-E. Rather, doubling of *acute 1* plasma GFAP was significantly associated with higher odds of having a worse *6m* GOS-E (OR=1.31, p=0.0003) and *12m* GOS-E (OR=1.46, p<0.0001).

### Different trajectories of plasma VEGF-A are associated with different clinical features

We investigated whether trajectory of plasma VEGF-A had clinical relevance. In the subset of BIO-AX-TBI TBI participants where all *acute 1, acute 2* and *10d6w* samples were available (n=118), we split participants into four trajectory categories based on VEGF-A behaviour over these timepoints: peak (levels initially climb, then decreased), trough (levels initially fall, then increase), rising (levels increase from the start), falling (levels fall from the start) (FIG 5A). TBI patients with a rising VEGF-A trajectory had significantly lower subacute (*10d6w*) white matter integrity in both corpus callosum (coefficient= –1.37, *p=0.013*) and whole skeleton (coefficient= –0.54, *p=0.012*), as compared with patients with a falling trajectory (FIG 5B, C). Trajectory type was not associated with demographic or clinical characteristics (SI TABLE5).

**Figure 5:**
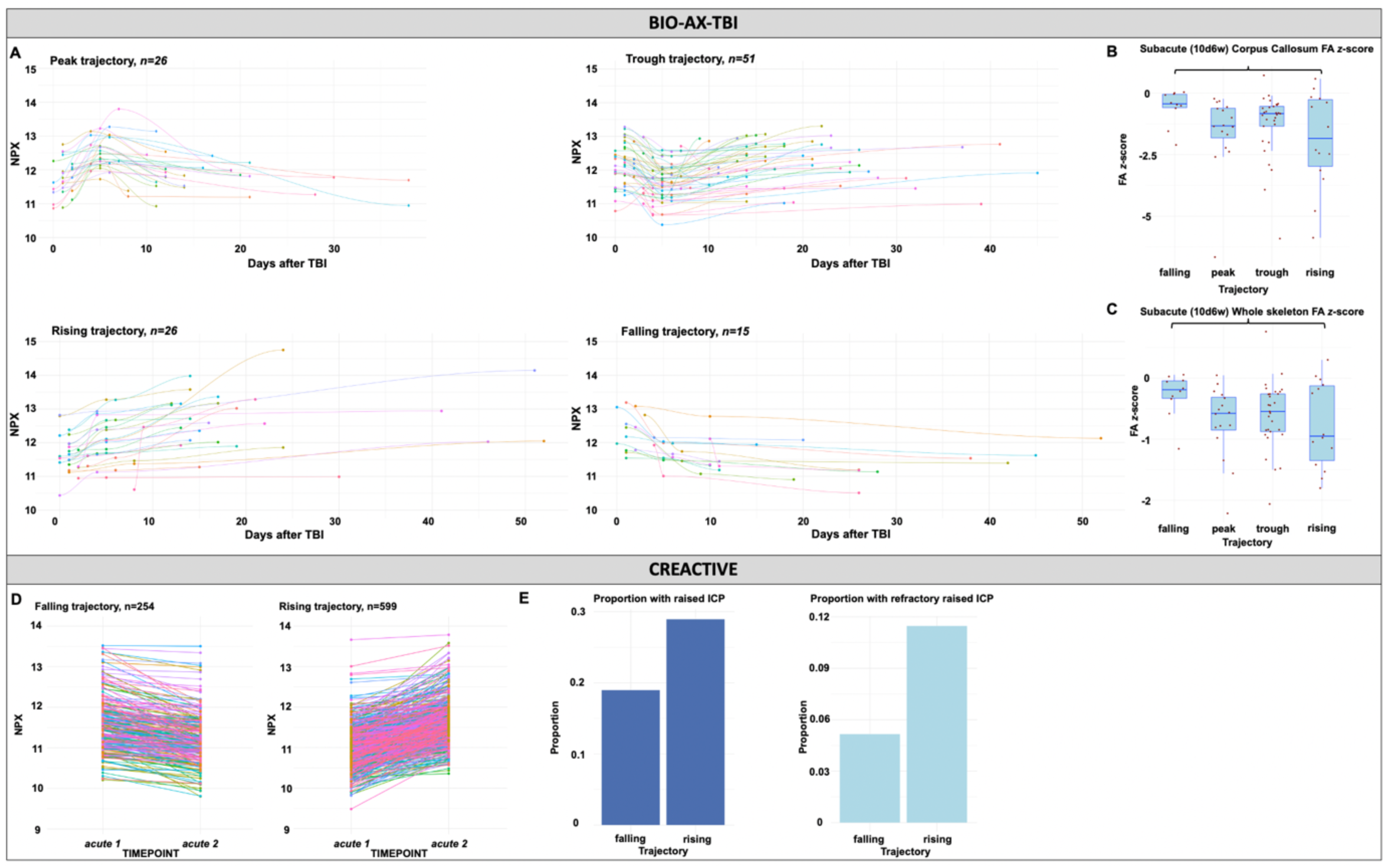
The BIO-AX-TBI cohort (A) split into 4 VEGF-A trajectory categories, based on change in levels over the acute-subacute period, illustrated by individual LOESS curves. (B) Corpus callosum FA z-score and (C) whole skeleton mean FA z-score in the subacute (10 days to 6 weeks, 10d6w) period in different trajectory sub-groups. Brackets indicate statistically significant comparison. The CREACTIVE cohort (D) split into 2 VEGF-A trajectory categories based on change in levels between the two acute timepoints. (E) The proportion of participants in each trajectory category which had raised intracranial pressure (ICP) and refractory raised ICP.

In the CREACTIVE cohort where both acute samples were available (n=853), we split participants into 2 trajectory categories, defined by whether VEGF-A rose or fell between timepoints (FIG 5D). A higher proportion of patients with a rising trajectory, as compared with a falling VEGF-A trajectory, had a pre-hospital GCS of 3-8 (48% v 38%, Ξ^2^(2)=7.05, p=0.03), raised ICP (OR=2.07, p=0.0016) and refractory raised ICP (OR=2.43, p=0.01). Trajectory type was not associated with GOS-E (SI TABLE6).

## DISCUSSION

We characterise plasma VEGF-A expression after traumatic brain injury (TBI) and explore its clinical associations, replicating findings across two multi-centre cohorts. We confirm previous reports of elevated plasma VEGF-A acutely after TBI and additionally show that, while this increase is not TBI-specific, the trajectory of plasma VEGF-A distinguishes TBI from NTT. Associations between plasma VEGF-A and TBI clinical severity were inconsistent, but VEGF-A levels correlated with neuronal and astroglial TBI biomarkers. Higher acute VEGF-A was independently associated with increased odds of refractory raised intracranial pressure (r-rICP) but not intracranial haemorrhage. Plasma VEGF-A correlated with lesion volume, and a rising VEGF-A trajectory was associated with having rICP and more white matter injury. Finally, higher acute VEGF-A predicted worse functional outcomes, partly reflecting initial injury severity.

The non-specific rise in plasma VEGF-A early after both TBI and non-TBI trauma is in keeping with the wide expression of VEGF-A and its multiplicity of functions, such as angiogenesis, bone formation and wound healing^2,31,32^. However, the later trajectory of plasma VEGF-A in TBI diverges from NTT diverge, in whom VEGF-A levels fall over the first few days post-injury. This suggests that early acute plasma VEGF-A (up to ∼5 days) non-specifically reflect injury, whilst later levels reflects TBI-specific pathophysiology.

Our findings are in line with a previous clinical TBI study showing plasma VEGF-A peaking at day 14^7^. This trajectory contrasts with many neuronal and astroglial TBI biomarkers, which peak within hours to days, and then drop rapidly. We also demonstrated that VEGF-A trajectory was associated with amount of subacute white matter injury, and occurrence of rICP/ refractory rICP. This supports VEGF-A being involved in secondary injury processes that get reflected as sustained high or rising plasma levels. For example, VEGF-A released from monocytes and astrocytes early after TBI promotes neuroinflammation, increased BBB permeability and recruitment of inflammatory cells^3,5,7,10,33,34^. An individual’s VEGF-A trajectory may be more informative about resulting damage than one-off measurements, a key consideration when considering timing of therapies targeting VEGF-A.

Increased plasma VEGF-A in acute TBI does not seem to be wholly explained by initial TBI severity. Higher acute plasma VEGF-A levels were associated with greater clinical severity in the BIO-AX-TBI cohort, but not the CREACTIVE cohort. The CREACTIVE cohort could be more severe, given their recruitment from ICU, suggesting a potential ‘ceiling’ for the relationship between plasma VEGF-A and injury severity.

There were relationships between acute and subacute plasma VEGF-A with acute and subacute neuronal/ astroglial TBI biomarkers, most consistently with NFL and UCH-L1, and with chronic NFL and lesion volumes. However, there were no relationships with MRI measures of white or grey matter atrophy. Therefore, acute and subacute plasma VEGF-A levels seem to be most involved with processes determining resulting extent of injury, but not those processes leading to ongoing neurodegeneration.

VEGF-A expression is seen in human TBI oedematous tissue, and involved in post-TBI BBB breakdown in animal TBI models^3,10,35^. Additionally, VEGF-A is involved in altered cerebrovascular reactivity and autoregulation^36,37^, which clinical studies demonstrate is associated with development of oedema, a risk for raised ICP^38^. Therefore, our finding, that higher late acute (∼5-10 days) plasma VEGF-A levels is independently associated with increased odds of refractory raised intracranial pressure (r-rICP), fit with the idea that VEGF-

A contributes to r-rICP through effects on BBB permeability or other cerebrovascular dysfunction^3,5,10,33^. Our finding that early acute GFAP is also associated with rICP/ r-rICP fits with the literature showing that greater injury severity increases risk of rICP/r-rICP^39^. The lack of association between plasma VEGF-A and presence of intracranial haemorrhage, plus a previous finding of lower microdialysate VEGF-A levels in subarachnoid haemorrhage compared to TBI^9^, suggests that plasma VEGF-A does not simply reflect vascular injury itself.

Previous work showed time-varying relationships between VEGF-A levels and functional outcomes – high levels early post-TBI (day 7) were associated with poor outcomes, whilst high subacute (day 21) levels were associated with better outcomes^7^. We partially replicated this, finding that higher early acute VEGF-A levels in the CREACTIVE cohort (majority of samples <7 days post-TBI) were associated with worse *6m* functional outcomes, independent of age and injury severity. The difference in VEGF-A levels was most marked between the worst (DEADVS) and other outcome categories, indicating that VEGF-A may be most useful for prognosticating extremes of outcome. In the BIO-AX-TBI cohort, no significant association was found after adjustment for initial injury severity, which may reflect lower statistical power or that VEGF-A is more discriminative in more severely injured cohorts like CREACTIVE.

### Limitations

Our study reveals novel clinical associations of plasma VEGF-A after TBI. However, we cannot conclude about mechanisms or causality. Exact timing information for when rICP/r-rICP developed is unavailable, so we cannot conclude whether plasma VEGF-A is a contributor or bystander in this pathology. Future mechanistic studies should investigate the exact role of VEGF-A in post-TBI secondary injury, and its interactions with associated proteins. For example, VEGI (vascular endothelial growth inhibitor) reduces inflammation and BBB disruption in experimental TBI^5^. While our findings suggest that higher acute VEGF-A is clinically detrimental, animal studies have not consistently shown that blocking VEGF-A improves outcomes, which may reflect the multiplicity and time-varying nature of its actions^40–42^. For example, VEGF receptor 1 is expressed predominantly on neurons, whereas VEGF receptor 2 is expressed more widely ^43^, and the two receptors have differential effects during angiogenesis^44^. Therefore, it is crucial to elucidate the specific pathways and timings for VEGF-A’s effects, which may also vary across patients.

## Conclusions

We analysed data across two clinical TBI cohorts to characterise the nature, trajectory and clinical associations of post-TBI plasma VEGF-A expression. We show independent associations between high plasma VEGF-A and refractory rICP, brain injury extent and functional outcome. Future studies should aim to disentangle the role of VEGF-A at different timepoints and specific pathways for its effects on outcomes after injury.

## Acknowledgements

ERA-NET NEURON Cofund (MR/R004528/1), a part of the European Research Projects on External Insults to the Nervous System call, within the Horizon 2020 funding framework, and the European Union’s Seventh Framework Programme (FP7/2007-2013, Grant Agreement number 602714) provided the core funds for the project. The UK Dementia Research Institute provided additional funds. LML and NG are supported by NIHR academic clinical lectureships, and acknowledges the support of the Imperial NIHR BRC. NG acknowledges support of Academy of Medical Sciences. HZ is a Wallenberg Scholar and a Distinguished Professor at the Swedish Research Council supported by grants from the Swedish Research Council (#2023-00356; #2022-01018 and #2019-02397), the European Union’s Horizon Europe research and innovation programme under grant agreement No 101053962, Swedish State Support for Clinical Research (#ALFGBG-71320), the Alzheimer Drug Discovery Foundation (ADDF), USA (#201809-2016862), the AD Strategic Fund and the Alzheimer’s Association (#ADSF-21-831376-C, #ADSF-21-831381-C, #ADSF-21-831377-C, and #ADSF-24-1284328-C), the European Partnership on Metrology, co-financed from the European Union’s Horizon Europe Research and Innovation Programme and by the Participating States (NEuroBioStand, #22HLT07), the Bluefield Project, Cure Alzheimer’s Fund, the Olav Thon Foundation, the Erling-Persson Family Foundation, Familjen Rönströms Stiftelse, Stiftelsen för Gamla Tjänarinnor, Hjärnfonden, Sweden (#FO2022-0270), the European Union’s Horizon 2020 research and innovation programme under the Marie Skłodowska-Curie grant agreement No 860197 (MIRIADE), the European Union Joint Programme – Neurodegenerative Disease Research (JPND2021-00694), the National Institute for Health and Care Research University College London Hospitals Biomedical Research Centre, and the UK Dementia Research Institute at UCL (UKDRI-1003).

