## Supplementary for "Acute plasma VEGF-A levels are associated with raised intracranial pressure and chronic lesion volume after Traumatic Brain Injury"

**Supplementary Information**

**Supplementary Methods:**

*Associations between VEGF-A and neuronal/ astroglial TBI biomarkers*

We examined correlations at the *acute 1, acute 2* and, for BIO-AX-TBI, at the subacute (10d6w) timepoints and for peak values. Peak values for each individual were defined as the highest biomarker level across the *acute 1, acute 1* and *10d6w* timepoints. In the chronic setting, NFL (neurofilament light), total Tau and GFAP are thought to indicate ongoing neuronal injury or neurodegeneration^15,20,21^. Therefore, we also examined the correlation between peak VEGF-A and 6 and 12-month NFL, GFAP and total Tau. We used Spearman’s correlations with FDR correction for multiple comparisons across all correlations examined.

*Associations between VEGF-A and raised intracranial pressure, refractory raised intracranial pressure and intracranial haemorrhage*

Separate models were used to test associations between *acute 1* and *acute 2* VEGF-A levels and each outcome (ICH, rICP and r-rICP). In the BIO-AX-TBI cohort, Chi-squared tests were used to investigate whether subgroups of TBI patients with different VEGF-A trajectory types differed in proportions of patients who had ICH, rICP and r-rICP. Additionally, models were built to test whether VEGF-A trajectory type were associated with ICH, rICP and r-rICP, including the same covariates. The numbers and demographics of TBI patients going into each analysis, based on data availability, are summarised in Supplementary Table 1.

**Supplementary Table 1**: Numbers and demographics of TBI patients included in each linear model examining relationship between plasma VEGF-A and raised intracranial pressure, refractory raised intracranial pressure and intracranial haemorrhage

| *Independent variable = VEGF-A trajectory type*  *Dataset = BIO-AX-TBI* | | | |
| --- | --- | --- | --- |
| *Model(s)* | *Numbers* | *Age*  Mean *(sd)* | *Sex* |
| rICP, r-rICP, ICH | 86 | 46.5 years  *(17.8 years)* | 12F:74M |
| *Independent variable = acute 1 VEGF-A*  *Dataset = BIO-AX-TBI* | | | |
| rICP, r-rICP, ICH | 138 | 45.0 years  *(17.4 years)* | 29F:109M |
| *Independent variable = acute 2 VEGF-A*  *Dataset = BIO-AX-TBI* | | | |
| rICP, r-rICP, ICH | 120 | 46.1 years  *(18.0 years)* | 22F:98M |
| *Independent variable = acute 1 VEGF-A*  *Dataset = CREACTIVE* | | | |
| ICH | 934 | 55.5 years  *(20.4 years)* | 226F:708M |
| rICP, r-rICP | 932 | 55.4 years  *(20.4 years)* | 226F:706M |
| *Independent variable = acute 2 VEGF-A*  *Dataset = CREACTIVE* | | | |
| ICH | 709 | 56.5 years  *(20.1 years)* | 168F:541M |
| rICP, r-rICP | 707 | 56.4 years  *(20.0 years)* | 168F:539M |

rICP=raised intracranial pressure; r-rICP=refractory raised intracranial pressure; ICH=intracranial haemorrhage

*Associations between VEGF-A and MRI measures of injury*

We investigated the following relationships with peak VEGF-A: change in grey matter volume between 10d6w and 6 months (GM JD 10d6w-6m), and change in white matter volume between 6 and 12 months (WM JD 6m-12m) (2 imaging measures previously found to be different between the TBI and CON cohorts of BIO-AX-TBI^14^), whole skeleton mean fractional anisotropy (FA) *z-*score at 10d6w, 6 and 12 months, corpus callosum mean FA at 10d6w, 6 and 12 months, and lesion volume at 10d6w, 6 and 12 months.

The linear models had the following covariates, informed by the literature:

- GM JD 10d6w-6m model: age, peak values of GFAP, NFL, TAU, UCH-L1 and S100B (these biomarkers have previously been shown to be significantly associated with GM JD 10d6w-6m^14^)
- WM JD 6m-12m model: age, peak NFL (peak NFL has previously been shown to be significantly associated with WM JD 6m-12m)
- Whole skeleton and corpus callosum mean FA models: age, peak values of NFL and TAU (these biomarkers have previously shown significant associations with FA^14^)
- Lesion volume models: age, *acute 1* GFAP

Values of GFAP, NFL, Tau, UCH-L1 and S100B were log2 transformed to be more comparable to the VEGF-A NPX unit. Where there was a significant association between peak VEGF-A level and the neuroimaging measure, Spearman’s correlations were calculated to illustrate the direction and strength of bivariate correlation between peak VEGF-A levels and the neuroimaging measure. The numbers and demographics of TBI patients going into each analysis, based on data availability, are summarised in Supplementary Table 2. Additionally, models were built to test whether VEGF-A trajectory type was associated with lesion volume, and whole skeleton and corpus callosum FA *z*-score at 10d6w, using the same covariates.

**Supplementary Table 2:** Numbers and demographics of TBI patients included in each linear model examining relationship between VEGF-A and MRI measures, in the BIO-AX-TBI cohort

| *Independent variable = peak VEGF-A* | | | |
| --- | --- | --- | --- |
| *Model(s)* | *Numbers* | *Age*  Mean *(sd)* | *Sex* |
| Change in grey matter volume between 10d6w and 6 months  (GM JD 10d6w-6m) | 56 | 45.2 years  *(16.2 years)* | 11F:45M |
| Change in white matter volume between 6 and 12 months  (WM JD 6m-12m) | 36 | 48.4 years  *(16.0 years)* | 9F:27M |
| Whole skeleton and Corpus Callosum mean fractional anisotropy *z-*score at 10d6w | 125 | 44.8 years  *(17.1 years)* | 27F:98M |
| Whole skeleton and Corpus Callosum mean fractional anisotropy *z-*score at 6m | 50 | 46.4 years  *(15.7 years)* | 12F:38M |
| Whole skeleton and Corpus Callosum mean fractional anisotropy *z-*score at 12m | 37 | 49.1 years  *(15.6 years)* | 9F:28M |
| Lesion volume at 10d6w | 114 | 45.0 years  *(17.7 years)* | 22F:92M |
| Lesion volume at 6m | 46 | 45.2 years  *(16.9 years)* | 11F:35M |
| Lesion volume at 12m | 33 | 48.6 years  *(15.4 years)* | 9F:24M |
| *Independent variable = VEGF-A trajectory type* | | | |
| Whole skeleton and Corpus Callosum mean fractional anisotropy *z-*score at 10d6w | 69 | 46.3years  *(18.1 years)* | 11F:58M |
| Lesion volume at 10d6w | 69 | 45.8 years  *(17.9 years)* | 7F:62M |

Only models to test associations between VEGF-A trajectory type and subacute (10d6w timepoint) whole skeleton mean FA z-score, corpus callosum mean FA z-score and lesion volume were built. There were fewer than 30 participants’ worth of data available for models testing association between trajectory type and other possible neuroimaging measures, so these were not tested due to high risk of overfitting the model.

10d6w = 10 days to 6 weeks after injury; 6m = 6 months; 12m = 12 months

*Associations between VEGF-A and functional outcome*

Models were built to test associations between *acute 1* and *acute 2* VEGF-A and 6-month GOS-E in CREACTIVE dataset, and to test associations between peak VEGF-A and VEGF-A trajectory type and 6 and 12-month GOS-E in BIO-AX-TBI dataset.

**Supplementary Table 3**: Numbers and demographics of TBI patients included in each linear model examining relationship between plasma VEGF-A and functional outcome

| *Independent variable = VEGF-A trajectory type*  *Dataset = BIO-AX-TBI* | | | |
| --- | --- | --- | --- |
| *Model(s)* | *Numbers* | *Age*  Mean *(sd)* | *Sex* |
| 6 month GOS-E | 71 | 45.5 years  *(17.7 years)* | 9F:62M |
| 12 month GOS-E | 66 | 45.9 years  *(18.4 years)* | 10F:56M |
| *Independent variable = peak VEGF-A*  *Dataset = BIO-AX-TBI* | | | |
| 6 month GOS-E | 121 | 44.8 years  *(17.8 years)* | 26F:95M |
| 12 month GOS-E | 106 | 45.9 years  *(18.1 years)* | 26F:80M |
| *Independent variable = acute 1 VEGF-A*  *Dataset = CREACTIVE* | | | |
| 6 month GOS-E | 910 | 55.3 years  *(20.4 years)* | 223F:687M |
| *Independent variable = acute 2 VEGF-A*  *Dataset = CREACTIVE* | | | |
| 6 month GOS-E | 691 | 56.4 years  *(20.1 years)* | 167F:524M |

GOS-E = Glasgow Outcome Scale Extended

**Supplementary Results**

*Associations of plasma VEGF-A with age and presence of extracranial injury*

**Supplementary Figure 1**

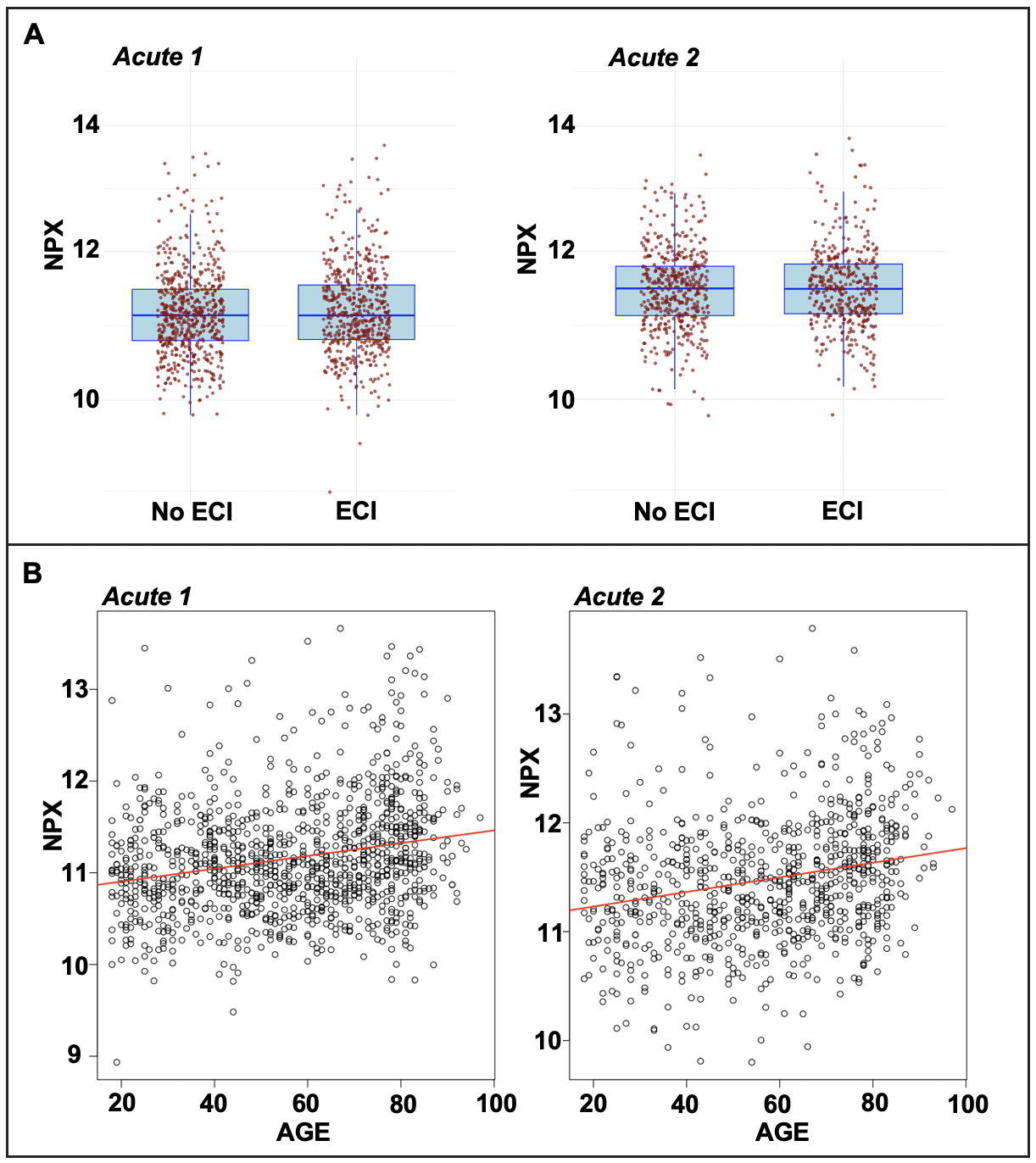

*SI FIGURE 1:*  In the CREACTIVE cohort: (A) Plasma VEGF-A levels in TBI patients with and without additional extracranial injury (ECI) at the *acute 1* and *acute 2* timepoints. (B) Relationship between plasma VEGF-A levels and age at the *acute 1* and *acute 2* timepoints.

**Supplementary Figure 2**

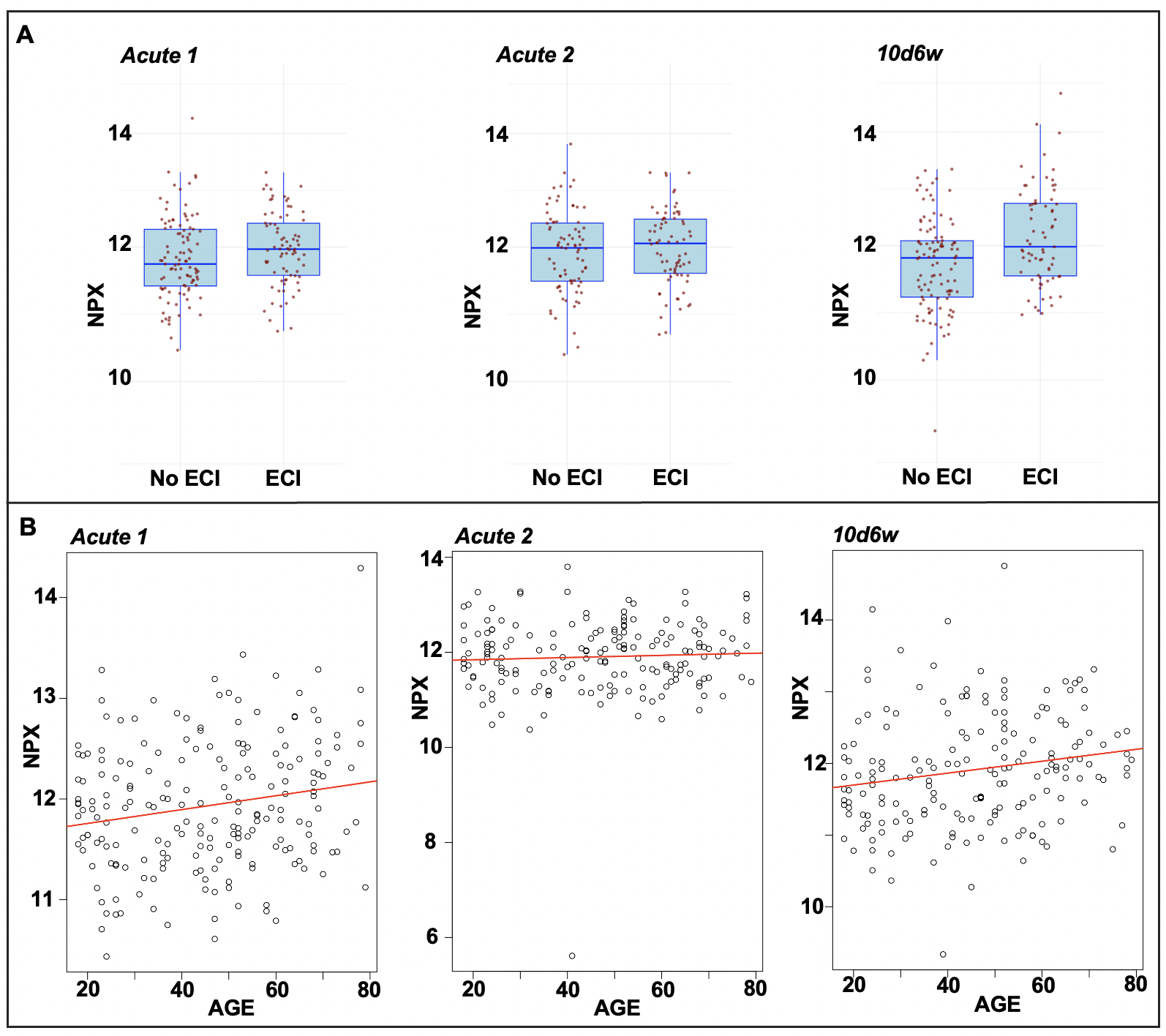

*SI FIGURE 2*: In the BIO-AX-TBI cohort: (A) Plasma VEGF-A levels in TBI patients with and without additional extracranial injury (ECI) at the *acute 1*, *acute 2* and *10d6w* timepoints. (B) Relationship between plasma VEGF-A levels and age at the *acute 1, acute 2* and *10d6w* timepoints.

*Associations between acute 1 GFAP, age and rICP, rICP and between VEGF-A and ICH.*

In the CREACTIVE data, there was a significant positive association of *acute 1* plasma GFAP with occurrence of both rICP and r-rICP. For a doubling of GFAP, the odds of having rICP was 1.56 (p<0.0001) in the *acute 1* VEGF-A model, and 1.46 (p<0.0001) in the *acute 2* VEGF-A model; the odds of having r-rICP was 1.60 (p<0.0001) in the *acute 1* VEGF-A model, and 1.64 (p<0.0001) in the *acute 2* VEGF-A model. There was a significant negative association of age with occurrence of both rICP and r-rICP. For a 1-year increase in age, the odds of having rICP was 0.97 (p<0.0001) in the *acute 1* VEGF-A model, and 0.97 (p<0.0001) in the *acute 2* VEGF-A model; the odds of having r-rICP was 0.98 (p=0.0002) in the *acute 1* VEGF-A model, and 0.96 (p<0.0001) in the *acute 2* VEGF-A model. There was no association between acute plasma VEGF-A and presence of ICH (FIG3C). However, there was a positive association between *acute 1* GFAP and presence of ICH in the *acute 1* VEGF-A model, with odds of 1.09 (p=0.018) of ICH for a 1-unit increase in GFAP. There was also a positive association between age and presence of ICH. For a 1-year increase in age, the odds of having ICH on admission CT was 1.04 (p<0.0001) in the *acute 1* VEGF-A model, and 1.04 (p<0.0001) in the *acute 2* VEGF-A model.

In the BIO-AX-TBI data, there was also a significant positive association between *acute 1* GFAP and probability of r-rICP, with an odds of 1.73 (p=0.0003) of r-rICP for a doubling of plasma GFAP, in the model with *acute 1* plasma VEGF-A levels only. There was a significant negative association between age and probability of both rICP and r-rICP. The odds of developing rICP for a 1-year increase in age was 0.96 (p=0.0005) in the model with *acute 1* VEGF-A, and 0.96 (p=0.0003) in the model with *acute 2* VEGF-A. The odds of developing r-rICP for a 1-year increase in age was 0.94 (p=0.0004) in the model with *acute 1* VEGF-A, and 0.94 (p=0.001) in the model with *acute 2* VEGF-A. There were no significant association between *acute 1* and *acute* 2 plasma VEGF-A level and probability of having intracranial haemorrhage on the initial CT brain scan (FIG5F).

*Relationship between VEGF-A and neuronal/ astroglial TBI biomarkers*

In the CREACTIVE cohort, *acute 1* VEGF-A levels (D0) showed a small positive correlation only with plasma NFL levels (r_s_=0.23, FDR-corrected p<0.05). *acute 2* levels of VEGF-A were moderately correlated with *acute 2* NFL, GFAP, total TAU and UCH-L1 (r_s_=0.27, r_s_=0.20, r_s_=0.24, r_s_=0.26 respectively, FDR-corrected p<0.05 for all) (SI FIG3B). In the BIO-AX-TBI cohort, *acute* *1* plasma VEGF-A showed small positive correlations with NFL (r_s_=0.18), total TAU (r_s_=0.2), UCH-L1 (r_s_=0.27) and S100B (r_s_=0.24, all FDR-corrected <0.05). *acute 2* VEGF-A only showed a small positive correlation with UCH-L1 (r_s_=0.22, FDR-corrected p<0.01). At the subacute timepoint (*10d6w*), VEGF-A levels showed small to moderate positive correlations with NFL (r_s_=0.34), GFAP (r_s_=0.38), UCH-L1 (r_s_=0.43) and S100B (r_s_=0.18, all FDR-corrected p<0.05) (SI FIG3D).

Given that we found the peak of plasma VEGF-A in the BIO-AX-TBI cohort to be at 16 days, and that there was spread of days on which samples were taken across the acute and subacute timepoints, we also investigated the relationship of biomarkers with peak plasma VEGF-A level in the BIO-AX-cohort. We found that peak plasma VEGF-A showed small to moderate positive correlations with peak levels of NFL (r_s_=0.29), GFAP (r_s_=0.29), total TAU (r_s_=0.30), UCH-L1 (r_s_=0.39) and S100B (r_s_=0.41, all FDR-corrected p<0.05). Further, we found that peak VEGF-A levels had a moderate positive correlation with plasma NFL levels at 6 months (r_s_=0.41, FDR-corrected p<0.05) (SI FIG3D). There were no significant correlations between peak VEGF-A and neuronal/ astroglial marker levels at 12 months.

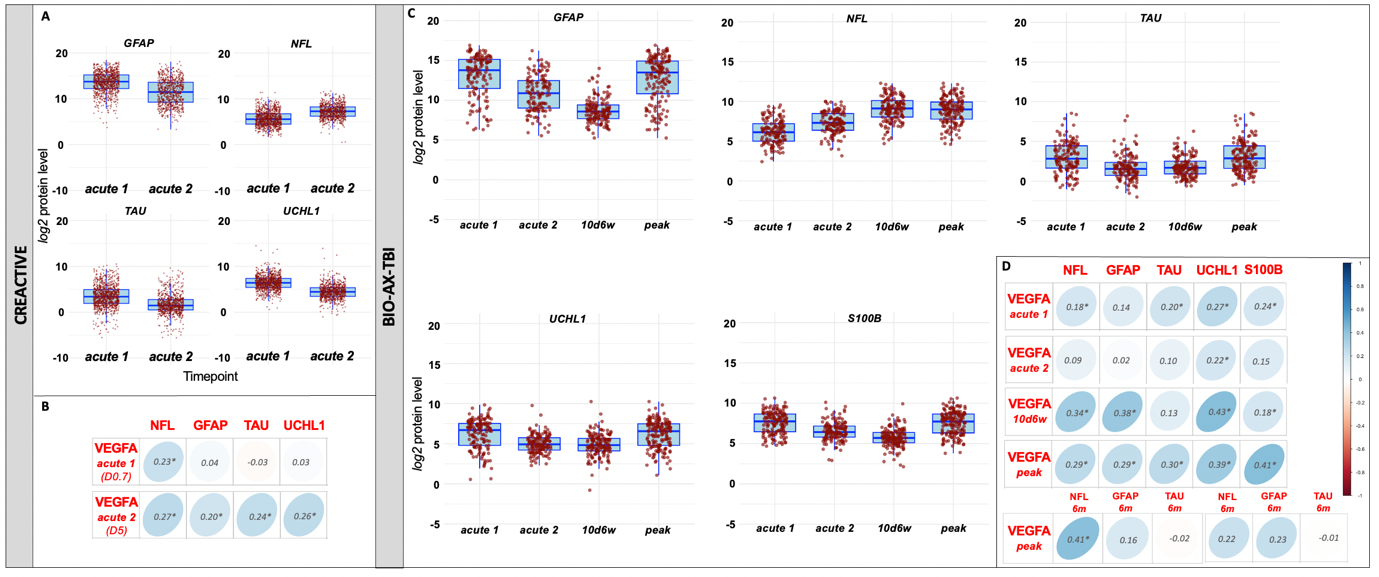

***Supplementary Figure 3:*** *For the CREACTIVE cohort:* *(A) Neuronal/ astroglial marker plasma levels at acute 1 and acute 2 timepoints (B) Correlations between plasma VEGF-A levels and neuronal and astroglial biomarkers at each timepoint. (B) Correlations between plasma VEGF-A levels and neuronal and astroglial biomarkers at the acute 1, acute 2 and subacute (10 days to 6 weeks, 10d6w) timepoints in the BIO-AX-TBI cohort. Mean sampling day shown in brackets (e.g. D23). For the BIO-AX-TBI cohort: (C) Neuronal/ astroglial marker plasma levels at acute 1, acute 2 and subacute (10d6w) timepoints for the TBI patients (D) Correlations between plasma VEGF-A levels and neuronal/astroglial biomarkers at each timepoint and peak plasma VEGF-A level and 6 month (6m) neuronal/astroglial biomarkers. Peak VEGF-A level defined as the highest value from acute 1, acute 2 and subacute timepoints. Mean sampling day shown in brackets (e.g. D5). Correlation values shown are Spearman’s rho correlation coefficients, and *denotes correlations which are statistically significant after FDR correction. NFL = neurofilament light, GFAP = glial fibrillary acidic protein, TAU = total Tau, UCHL1 = ubiquitin C-terminal hydrolase 1, S100B = S100 calcium binding protein B.*

*Association between plasma VEGF-A and MRI measures of injury*

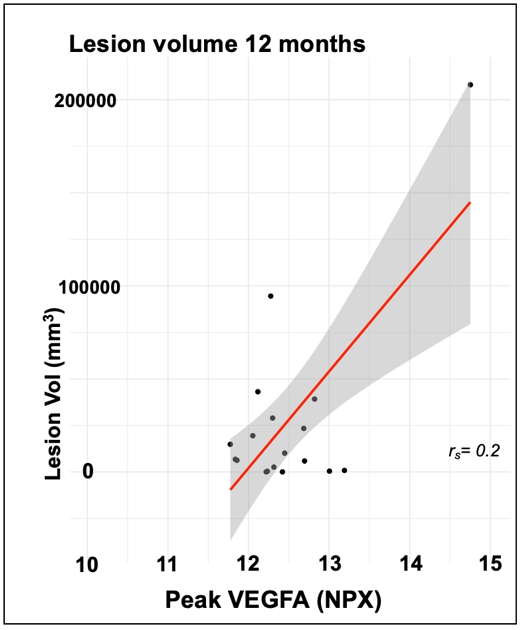

***Supplementary Figure 4:*** *Bivariate correlations between peak plasma VEGF-A levels (from day of TBI up to 6 weeks after TBI) with lesion volume at 12 months. Correlation value is Spearman’s rho coefficients (r_s_).*

*Acute 1* GFAP level was not significantly associated with *12m* lesion volume (coefficient=2226.2, p=0.37), but was significantly associated with *10d6w* lesion volume (coefficient=9644.8, p=0.02) when peak VEGF-A was not (coefficient=27,514.0, p=0.09). There were no other significant associations between peak plasma VEGF-A and MRI measures (either static or dynamic), after accounting for covariates.

*Association between plasma VEGF-A and functional outcome*

**Supplementary Table 4**: summary of Dunn’s test statistic and significance level for all significant post-hoc comparisons

| ***CREACTIVE*** | **Comparison** | | **Dunn’s test *z* statistic** | ***p*** |
| --- | --- | --- | --- | --- |
| ***acute 1* VEGF-A** | DEADVS | POOR | 5.11 | <0.0001 |
|  | DEADVS | MOD | 5.02 | <0.0001 |
|  | DEADVS | GOOD | 8.06 | <0.0001 |
|  | MOD | GOOD | 3.52 | 0.001 |
| ***acute 2 VEGF-A*** | DEADVS | POOR | 7.12 | <0.0001 |
|  | DEADVS | MOD | 7.23 | <0.0001 |
|  | DEADVS | GOOD | 9.31 | <0.0001 |
|  | MOD | GOOD | 3.35 | 0.002 |
| ***BIO-AX-TBI*** | **Comparison** | | **Dunn’s test *z* statistic** | ***p*** |
| **peak VEGF-A (6 months)** | DEADVS | GOOD | 2.42 | 0.046 |
|  | POOR | GOOD | 3.83 | 0.0004 |
|  | POOR | MOD | 3.83 | 0.002 |
| **Peak VEGF-A**  **(12 months)** | DEADVS | GOOD | 2.56 | 0.03 |
|  | POOR | GOOD | 2.75 | 0.02 |

In the CREACTIVE dataset, when adjusting for age and *acute 1* plasma GFAP, as a measure of extent of initial intracranial injury, a doubling of *acute 1* and *acute 2* VEGF-A levels corresponded with increased odds of moving to a worse GOS-E category (doubling of *acute 1* VEGF-A OR=1.72, p<0.0001, doubling of *acute 2* VEGF-A OR=2.51, p<0.001). There was also a significant, but smaller, association of age and *acute 1* GFAP levels with 6-month outcome. A doubling of *acute 1* plasma GFAP was associated with an OR of moving to a worse GOS-E category of 1.25 (p<0.001) for the *acute 1* VEGF-A model and an OR of 1.21 (p<0.0001) for the *acute 2* VEGF-A model. Similarly, a 1-year increase in age was associated with an increased odds of moving to a worse GOS-E category in both *acute 1* VEGF-A (OR=1.03, p<0.0001) and *acute 2* (OR=1.03, p<0.0001) VEGF-A models.

*Associations of different VEGF-A trajectories*

**Supplementary Table 5**: Differences between subgroups within the BIO-AX-TBI cohort based on VEGF-A trajectory during the acute-subacute period

|  | **Mean age (y)** | ***s.d.*** |
| --- | --- | --- |
| **Peak** | 46.2 | *20.9* |
| **Trough** | 46.9 | *16.9* |
| **Rise** | 46.0 | *16.1* |
| **Fall** | 49.6 | *17.9* |
| *F* | 0.1539 *(p=0.927)* | |

|  | **Glasgow Coma Scale** | | |
| --- | --- | --- | --- |
|  | ***3-8*** | ***9-13*** | ***14-15*** |
| **Peak** | 17 (65%) | 3 (12%) | 6 (23%) |
| **Trough** | 32 (63%) | 11 (22%) | 8 (16%) |
| **Rise** | 15 (58%) | 7 (27%) | 4 (15%) |
| **Fall** | 7 (47%) | 6 (40%) | 2 (13%) |
| *X^2^* | *5.0391 (p=0.5388)* | | |

|  | **Present** | **Not present** |  |  |
| --- | --- | --- | --- | --- |
|  | ***Extracranial Injury*** | |  |  |
| **Peak** | 13 (50%) | 13 (50%) |  |  |
| **Trough** | 32 (63%) | 19 (37%) |  |  |
| **Rise** | 13 (50%) | 13 (50%) |  |  |
| **Fall** | 4 (27%) | 11 (73%) |  |  |
| *X^2^* | *6.29 (p=0.098)* | |  |  |
|  | ***Refractory Raised ICP*** | |  |  |
| **Peak** | 8 (31%) | 18 (69%) |  |  |
| **Trough** | 6 (12%) | 45 (88%) |  |  |
| **Rise** | 4 (15%) | 22 (85%) |  |  |
| **Fall** | 0 (0%) | 15 (100%) |  |  |
| *X^2^* | *8.02 (p=0.046)* | |  |  |
|  | ***Any Raised ICP*** | |  |  |
| **Peak** | 14 (54%) | 12 (46%) |  |  |
| **Trough** | 9 (18%) | 42 (82%) |  |  |
| **Rise** | 10 (38%) | 16 (62%) |  |  |
| **Fall** | 4 (27%) | 11 (73%) |  |  |
| *X^2^* | 11.32 (*p=0.01)* | |  |  |
|  | ***Intracranial haemorrhage*** | |  |  |
| **Peak** | 3 (12%) | 23 (88%) |  |  |
| **Trough** | 11 (22%) | 40 (78%) |  |  |
| **Rise** | 1 (4%) | 25 (96%) |  |  |
| **Fall** | 5 (33%) | 10 (67%) |  |  |
| *X^2^* | 7.35 (*p=0.06)* | |  |  |
|  | ***6-month Good Outcome^1^*** | | ***12-month Good Outcome^1^*** | |
| **Peak** | 8 (31%) | 16 (69%) | 9 (43%) | 12 (57%) |
| **Trough** | 24 (60%) | 16 (40% | 25 (69%) | 11 (21%) |
| **Rise** | 10 (50%) | 10 (50%) | 9 (50%) | 9 (50%) |
| **Fall** | 7 (64%) | 4 (36%) | 8 (73%) | 3 (27%) |
| *X^2^* | 5.00 (*p=0.17)* | | 5.35 (*p=0.15)* | |

^1^ Good Outcome = Glasgow Outcome Scale-Extended score 5 to 8

**Supplementary Table 6:** Differences between subgroups within the CREACTIVE cohort based on VEGF-A trajectory during the acute-subacute period

|  | **Mean age (y)** | ***s.d.*** |
| --- | --- | --- |
| **Rise** | 57.0 | 20.1 |
| **Fall** | 56.0 | 19.6 |
| *t* | -0.698 *(p=0.49)* | |

|  | **Glasgow Coma Scale** | | |
| --- | --- | --- | --- |
|  | ***3-8*** | ***9-13*** | ***14-15*** |
| **Rise** | 287 *(48%)* | 174 *(29%)* | 138 *(23%)* |
| **Fall** | 97 (38%) | 84 *(33%)* | 73 *(29%)* |
| *X^2^* | 7.045 *(p=0.0295)* | | |

|  | **Present** | **Not present** |
| --- | --- | --- |
|  | ***Extracranial Injury*** | |
| **Rise** | 279 *(47%)* | 320 *(53%)* |
| **Fall** | 122 *(48%)* | 132 *(52%)* |
| *X^2^* | 0.098 *(p=0.75)* | |
|  | ***Refractory Raised ICP*** | |
| **Rise** | 83 *(14%)* | 515 *(86%)* |
| **Fall** | 17 *(7%)* | 236 *(93%)* |
| *X^2^* | 8.113 *(p=0.004)* | |
|  | ***Any Raised ICP*** | |
| **Rise** | 173 *(29%)* | 425 *(71%)* |
| **Fall** | 48 *(19%)* | 205 *(81%)* |
| *X^2^* | 8.66 *(p=0.003)* | |
|  | ***Intracranial haemorrhage*** | |
| **Rise** | 477 *(80%)* | 122 *(20%)* |
| **Fall** | 206 *(81%)* | 48 *(19%)* |
| *X^2^* | 0.158 *(p=0.69)* | |
|  | ***6-month Good Outcome^1^*** | |
| **Rise** | 188 *(32%)* | 401 *(68%)* |
| **Fall** | 91 *(37%)* | 154 *(63%)* |
| *X^2^* | 1.893 *(p=0.169)* | |

^1^ Good Outcome = Glasgow Outcome Scale-Extended score 5 to 8
